# Inequalities in financial burden of severe childhood illness on households in Lao PDR: A prospective cohort study

**DOI:** 10.1101/2025.05.28.25328517

**Authors:** Alicia Quach, Mayfong Mayxay, Laddaphone Bounvilay, Amphaivanh Thammavong, Toukta Bounkhoun, Chom Phaiphichit, Nar Kingkeooudom, Sommanikhone Phangmanixay, Phouthalavanh Souvannasing, Elizabeth A Ashley, Cattram Nguyen, Natalie Carvalho, Fiona M Russell

**Author notes:** Corresponding author (AQ). These authors contributed equally to this work (Natalie Carvalho, Fiona M Russell).

## Abstract

Lao People’s Democratic Republic (PDR) introduced a National Health Insurance (NHI) scheme in 2016 to all provinces except Vientiane Capital City. We describe the financial impact on households related to treatment of severe childhood illness at a hospital covered by NHI, and one without NHI.

We conducted a prospective cohort study (2022-2024) in Lao PDR of children aged one month to <15 years admitted with severe illness at two hospitals: Salavan Provincial Hospital (SPH) with NHI, and National Children’s Hospital (NCH) without NHI, with two-month follow-up post-discharge. Illness-related direct and indirect costs were collected. We calculated household out-of-pocket (OOP) costs, impoverishment and catastrophic health expenditure (CHE, >10% annual household expenditure) rates and analysed relative risk (RR) of CHE by socioeconomic status.

200 participants were recruited from each hospital with demographic differences observed between hospitals in urban residence (NCH 87.0%, SPH 14.5%), maternal education (primary level: NCH 95.9%, SPH 76.3%) and wealth status (wealthiest quintile: NCH 79.0%, SPH 20.5%). Median household OOP costs were higher at NCH (USD290.6 [IQR 206.9–422.9]) compared to SPH (USD92.4 [IQR 56.3–52.9]). Impoverishment at two months post-discharge was 0.5% (95%CI 0.0-3.0) at NCH and 10.2% (95%CI 6.2-15.4) at SPH. CHE rates were 34.5% (95%CI 27.9-41.1) at NCH and 26.0% (95%CI 19.9-32.1) at SPH, with higher RR in the poorest versus wealthiest households (NCH: RR 6.6, 95%CI 4.5-9.5; SPH: RR 4.9, 95%CI 1.7-13.7) and households with no formal maternal education versus secondary education (NCH: RR 2.6, 95%CI 1.2-5.5; SPH: RR 4.6, 95%CI 1.92-11.1).

Direct medical costs were lower where NHI is available, but total household OOP costs and CHE rates were high at both hospitals, particularly among disadvantaged households. Additional interventions are required to prevent severe illness and provide financial protection for socioeconomically disadvantaged groups to reduce health-related economic burden on households in Lao PDR.

## Introduction

Under Sustainable Development Goal 3.8, the target by 2030, is to “achieve universal health coverage (UHC), including financial protection, access to quality essential health-care services and access to safe, effective, quality and affordable essential medicines and vaccines for all”.[1] Financial protection is the percentage of the population protected against catastrophic health expenditures (CHE) and healthcare related impoverishment. CHE captures households that incur out-of-pocket (OOP) payments exceeding a certain threshold of household expenditure or income, with thresholds used in the literature varying between 10% - 40%.[2–8] Impoverishment measures those pushed below poverty lines due to health related OOP payments.[2, 7, 9, 10] Most recent figures estimated that 13.5% of the world’s population experienced CHE and 4.4% were impoverished, based on the extreme poverty line, due to health care expenditures.[11] People living in low-and-middle-income-countries (LMICs) were almost exclusively affected, with higher rates of financial hardship occurring in the poor, those living in remote areas and marginalised groups.[11]

Health insurance is one strategy used to reduce OOP health costs, but coverage in LMICs remain low with variable impact on financial protection.[12] Health insurance tends to address direct medical costs such as investigations, medicines and hospital bed fees. However, OOP costs related to sick children requiring inpatient care, exceed the direct cost of medical treatment.[8, 10, 13] The additional burden of direct non-medical costs such as transportation and accommodation for the child and caregiver as well indirect costs from loss of productivity/income for caregivers who need to accompany their sick child, can create high financial distress on households. A systematic review evaluating OOP costs associated with treatment of childhood infectious diseases in LMICs found that direct non-medical OOP costs and indirect costs related to inpatient care, contributed 29% and 42% of total OOP costs, respectively.[10]

The Lao People’s Democratic Republic (Lao PDR), is a lower-middle income country in South-East Asia, which has a national poverty rate of 18.3%, with higher rates in rural and agricultural households, those where household heads had little or no formal education, and in minority ethnic groups.[14] Government spending on health care has remained relatively low compared to neighbouring countries, with OOP expenditure being the predominant source of health financing.[15, 16] Lao PDR has goals to attain UHC and has a particular focus on maternal and child health.[17] The National Health Insurance (NHI) scheme launched in 2016, aims to regulate user fees at health facilities and make quality health services more accessible to vulnerable populations.[15] The NHI was introduced into all provinces by 2019, excluding Vientiane Capital City, where residents continue to have access to the pre-existing insurance schemes. Under the NHI, essential health services are free for pregnant women, children under five years of age and poor households, with small co-payments for others. Poor households also receive reimbursements for transportation and other incidental costs, where barriers to access can impact health service uptake.[18]

Implementation of the NHI scheme into practice has been variable. Early studies following NHI introduction revealed variable OOP costs for inpatient and outpatient services with CHE rates reaching over 30%.[15, 18] To date, there have been no studies in Lao PDR since the introduction of NHI, evaluating levels of financial hardship for households with sick children requiring medical treatment. The primary aim of our study was to describe the financial impact (as measured by household OOP costs, CHE and impoverishment) of a severe childhood illness on households in the context of NHI, at hospitals with and without NHI. Our secondary aim was to evaluate the differences in financial burden across wealth quintile groups, geographical residence, maternal education and ethno-linguistic subpopulation groups. This will add to the data on financial protection and inequity gaps towards Lao PDR’s goal of achieving UHC.

## Methods

### Study design

This is a prospective cohort study of children admitted with severe illness to two different hospitals in Lao PDR, with community follow-up at two weeks and two months following discharge. Data were collected between 28 October 2022 and 10 January 2024 on health-related OOP costs from a household perspective.

### Study sites

Lao PDR has a population of about 7.6 million people, 15% under 15 years of age, mostly living in rural areas.[19] It is ethnically diverse with four formal ethno-linguistic groups (Lao-Tai, Mon-Khmer, Hmong Mien and Chinese-Tibetan), but over 200 self-identified ethnic subgroups.[20, 21] Lao PDR consists of 17 provinces and one prefecture, Vientiane Capital City. Nationally, there are eight central, tertiary level hospitals, 17 provincial, secondary level hospitals and 135 district hospitals.[16] Health insurance provided through the NHI scheme covers all the provinces, excluding Vientiane Capital City. Vientiane Capital, where about 13% of the country’s population live, is partially covered by social health protection schemes, including the free maternal and children under five policy. About 6% of the total population, all in Vientiane Capital, are not eligible for any form of health insurance.[22]

Participants were recruited from two hospitals: National Children’s Hospital (NCH), a central, tertiary level hospital with 70 paediatric beds in Vientiane Capital City not covered by NHI; and Salavan Provincial Hospital (SPH), a provincial, secondary level hospital with 49 paediatric beds in Salavan Province in Southern Lao PDR, fully covered by NHI. In 2023, NCH had 11,418 paediatric admissions and SPH had 3550 paediatric admissions. These study sites were selected by our Lao investigators, with the aim of representing two different regions of Lao PDR (central and south regions), and a wide spread of demographics, to reflect the Lao population. We also chose to recruit from the two higher hospital levels, with high admission rates to reach our target sample size.

### Study population

We enrolled children aged between one month of age and <15 years old, presenting to hospital with acute symptoms starting within 14 days prior to hospital admission, and diagnosed with a severe illness. A severe illness was defined as showing emergency signs as per WHO Pocketbook criteria and/or requiring hospital admission for further treatment.[23] Emergency signs included: obstructed or absent breathing, severe respiratory distress, central cyanosis, shock, coma, convulsions or severe dehydration.[23] We excluded neonates and those admitted to the neonatal intensive care unit (NICU), as they are a specific population that would likely skew results. We also excluded children admitted for planned procedures or treatment of chronic medical conditions to focus on OOP costs for acute severe medical events with unanticipated household costs. Parents/guardians of enrolled children were required to speak the Lao language and be contactable by phone.

### Study procedures

For each participant, four study visits were conducted – at enrolment (hospital admission), at hospital discharge, and then at two weeks and two months following hospital discharge. Informed consent procedures were conducted at enrolment. As all participants were under 18 years, written consent was obtained from a parent/legal guardian of the participant. Signed assent was also sought from participants age 12 – 15 years, assessed as physically well enough and having the mature capacity to understand the study. All study visits were conducted in-person with the parent/caregiver and child, except at two weeks post-discharge, where a phone interview was conducted. Those who could not attend in-person for the study visit at two months post-discharge, were contacted for a phone interview.

Study staff recorded data on paper-based questionnaires with the parent/caregiver and directly with the child, where appropriate. Information collected included household demographics and socioeconomic status (age of child, ethnicity, parental education levels and occupation, estimated income, housing materials and assets), household expenditures (general expenditures as well as OOP costs related to the child’s current illness), financial coping strategies (health insurance, savings, borrowing money, selling assets) and the child’s physical health outcome post-discharge (recovery status, death). Information on the child’s inpatient management (admitted ward, duration of stay, diagnosis and discharge destination) were collected from hospital medical records.

Household consumption data were collected as a recall of expenses over a two-week period, prior to enrolment, and prior to the two-week and two-month post-discharge study visits. A two-week period was used to minimise recall bias and align with the most recent Lao Expenditure Consumption Survey 6 (LECS6) methodology. Expenses collected included food, housing, utility bills, fuel, transportation, education, clothing, personal care, recreation, and medical expenditures for other household members. Data were used to estimate household expenditures over two months and over one year.

Data collected at each study visit on household OOP costs associated with the child’s current illness included: direct medical costs (medical consultation fees, diagnostics, medicines, hospital bed fees, hospital transfer costs and insurance co-pay if applicable); direct non-medical costs (travel costs for the patient and accompanying carers/household members, daily living expenses whilst in hospital such as food, accommodation, childcare costs not usually paid for other children); and indirect costs (estimated loss of income of the primary income earner and primary caregiver due to child’s illness). This was based on the caregiver reported estimated daily income for the primary income earner and primary caregiver and calculated as the sum of (reported daily income x days of productivity lost).

### Outcome definitions

#### Out-of-pocket costs

Total household OOP costs related to the severe illness were calculated by adding direct (medical and non-medical) costs less any reimbursements such as health insurance, with indirect costs, incurred from the onset of symptoms to two months post-discharge from hospital. OOP costs were calculated using the local currency, Lao Kip (LAK) and then converted to United States Dollars (USD) using the official World Bank exchange rate for 2023 of LAK 17689 = USD 1.[24]

#### Catastrophic health expenditures

A household was deemed to have CHE if OOP costs exceeded a certain percentage of household expenditures or capacity-to-pay, where capacity-to-pay was defined as the total household expenditures minus necessary subsistence expenditures (food, housing and utilities). We used four different thresholds to define CHE: 10% of two-monthly HE and 40% of two-monthly capacity-to-pay were used to assess short-term financial impacts; 10% of annual household expenditures and 40% of annual capacity-to-pay to assess longer-term financial impacts of OOP costs. We chose 10% of annual household expenditures to align with recommended indicators for SDG monitoring.[1, 2] 40% of annual capacity-to-pay was also used, as an alternative for CHE calculation deemed more sensitive at identifying poorer populations.[2, 4, 5, 25–27]. As our study only recruited children with acute illnesses, we included short-term financial impacts over two months, to reflect the acute nature of the illnesses.[2, 10, 26]

#### Impoverishment

Two definitions for impoverishment were used: the International Poverty Line set at $2.15/person/day at 2017 PPP and the Laos National Poverty Line set at LAK 280,910/person/month at 2019 local prices (USD 22.00/person/month), both adjusted for inflation.[11, 14] The international poverty line was used to align with SDG indicators and enable cross-country comparisons, and the national poverty line to be more representative of poverty within Lao PDR.[9, 11] Household expenditure was used as a proxy for household income in Lao PDR where households are mainly supported by farming and subsistence living and not formal salaries. A household was deemed impoverished if the household expenditure excluding the total direct (medical and non-medical) OOP costs, divided by the household headcount (converted to daily and monthly expenditures per person), was below the poverty line.

### Study size

The sample size was based on the primary objective of estimating average household OOP costs across both hospitals related to treatment of children with a severe illness in Lao PDR. We extended calculations to also allow comparison of OOP costs across wealth quintiles. A sample size of 60 participants per quintile was needed to obtain 90% power and a significance level of 0.05 to detect a mean difference of LAK 82,000 (USD 8.5) across successive wealth quintiles (based on population data from previous analyses of the Lao Expenditure Consumption Survey).[28] Accounting for 20% loss to follow up, and presumed equal distribution across the wealth quintiles, the total sample size was 375 across both hospitals.

### Data analysis

Participant demographics, primary clinical diagnoses, inpatient management and health outcomes were summarised by hospital. Categorical variables were summarised as frequencies and percentages of participants, and continuous variables with medians (IQR). Each household was assigned to a wealth quintile by first creating a wealth index score using Principal Components Analysis using 77 household demographics and consumption variables, where factor weights were equivalent to those used in the Laos Social Indicator Survey II (LSIS II).[29] Participants were then classified into wealth quintiles using thresholds for the national wealth quintiles.[29, 30]

For our primary aim, OOP costs, CHE and impoverishment rates were summarised by hospital. Medians (IQR) and means (SD) were calculated for OOP costs. Mean CHE and impoverishment percentage rates were calculated with 95% confidence intervals (CIs). We calculated impoverishment rates at baseline, two weeks and two months post-hospital discharge to determine if there were ongoing financial impacts from the severe illness.

For our secondary aim, OOP costs, CHE and impoverishment rates were disaggregated by wealth quintiles, maternal education categories, geographical residence and ethno-linguistic groups. Differences in median OOP costs across subgroups were estimated using quantile regression. Equity analyses included comparing risk ratios (RR) of CHE and impoverishment rates across the subpopulation groups, by hospital. Due to non-convergence, RR were estimated using Poisson regression with robust standard errors.

Data were entered and managed on REDCap electronic data capture software platform with manual second person data entry checks and inbuilt range checks for data cleaning prior to analysis. All statistical analyses were performed using statistical software package STATA (v18.0).[31]

### Ethical clearance

The study was conducted according to the protocol approved by The Royal Children’s Hospital Human Research Ethics Committee (HREC 81864/RCHM-2022), the University of Oxford Tropical Research Ethics Committee (552-22), and the Lao PDR Ministry of Health, National Ethics Committee for Health Research (2022.52).

## Results

Of the 474 children screened, a total of 400 participants were enrolled into the study, 200 from each hospital. Reasons for not being enrolled included parent decline, not having a telephone and not speaking the Lao language (Fig 1). Preliminary analysis after reaching the calculated sample size of 375 revealed participants were skewed towards the wealthier quintiles (Q4 and Q5). Recruitment therefore continued to try to reach the target of 60 participants for each wealth quintile, but was ceased before attaining this, deemed unlikely to be reached within a reasonable time period. 371 (92.8%) participants completed all four study visits with data available for complete case analysis.

**Figure 1.**
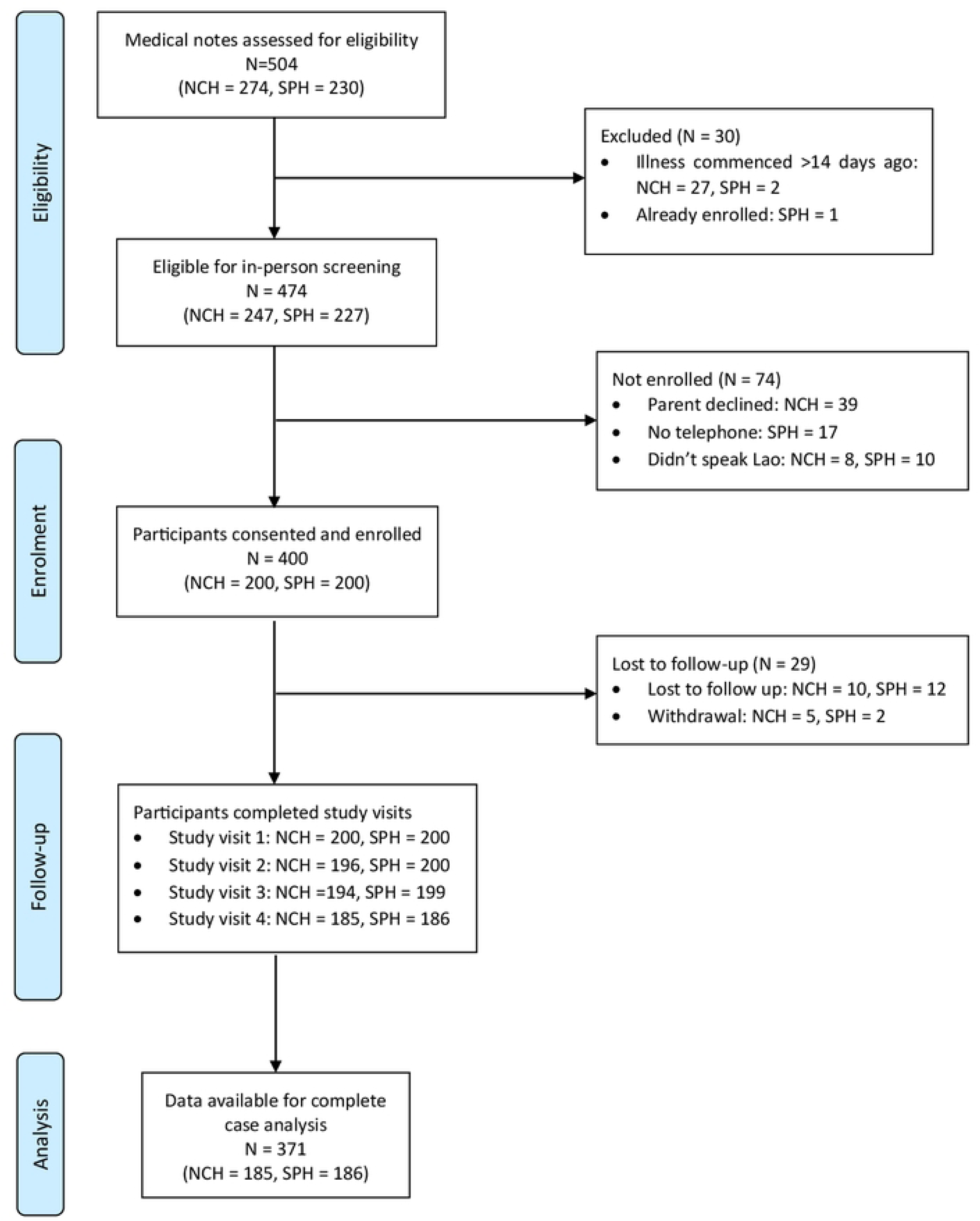
Study flowchart of participant recruitment and follow-up.

Table 1 summarises the demographic characteristics of participants and household members, by hospital. The median age of participants was one year (IQR 0.3 – 3.7 years) with most (82%) below five years of age. Characteristics differed between the two hospitals: almost all households (97.5%) from NCH were from the two wealthiest quintiles (Q4 and Q5), whereas households from SPH were more evenly distributed across wealth quintiles. Most households from NCH lived in urban areas (87%) whereas most from SPH lived in rural areas (85.5%). Mothers from NCH had higher education levels, with 85.1% having completed secondary level or higher, compared to SPH where 23.7% of mothers having had no formal education. All households from SPH were covered by the NHI. At NCH, 36.5% of households had some form of health insurance, mostly from the formal sector covering civil servants and private formal employees.

**Table 1.** Demographic characteristics of participants and household members.

|  | National Children's Hospital N (%) | Salavan Provincial Hospital N (%) |
| --- | --- | --- |
| Number of participants | N=200 | N=200 |
| Age of participant, in years |  |  |
| Median (IQR) | 0.9 (0.3, 2.4) | 1.4 (0.5, 4.9) |
| Age group |  |  |
| 1-11 months | 110 (55%) | 86 (43%) |
| 12-59 months | 66 (33%) | 65 (32.5%) |
| 60 months to <15 years | 24 (12%) | 49 (24.5%) |
| **Sex (participant)** |  |  |
| Male | 104 (52.0%) | 103 (51.5%) |
| Female | 96 (48.0%) | 97 (48.5%) |
| **Geographical residence** |  |  |
| Urban | 174 (87.0%) | 29 (14.5%) |
| Rural with road access | 26 (13.0%) | 171 (85.5%) |
| **Ethno-linguistic group** |  |  |
| Lao-Tai | 173 (86.5%) | 168 (84.0%) |
| Mon-Khmer | 6 (3.0%) | 29 (14.5%) |
| Hmong-Mien | 19 (9.5%) | 0 (0.0%) |
| Chinese-Tibetan | 1 (0.5%) | 0 (0.0%) |
| Other | 1 (0.5%) | 3 (1.5%) |
| **Household number** |  |  |
| Median (range) | 5 (3-14) | 5 (3-10) |
| **Relationship of respondent/guardian to participant** |  |  |
| Mother/stepmother | 160 (80.0%) | 179 (89.5%) |
| Father/stepfather | 33(16.5%) | 13 (6.5%) |
| Other | 7 (3.5%) | 8 (4.0%) |
| **Mother's education level** |  |  |
| None/early childhood | 8 (4.1%) | 45 (23.7%) |
| Primary | 21 (10.8%) | 62 (32.6%) |
| Secondary or higher | 165 (85.1%) | 83 (43.7%) |
| **Occupation of primary income earner** |  |  |
| Farmer/ agricultural | 9 (4.5%) | 126 (63.0%) |
| Manual laborer | 136 (68.0%) | 59 (29.5%) |
| Service and sales | 47 (23.5%) | 12 (6.0%) |
| Professional | 4 (2.0%) | 0 (0%) |
| House duties | 3 (1.5%) | 2 (1.0%) |
| Unemployed | 1 (0.50%) | 1 (0.5%) |
| **Wealth quintile (household)** |  |  |
| Q1 (poorest) | 0 (0.0%) | 21 (10.5%) |
| Q2 | 3 (1.5%) | 38 (19.0%) |
| Q3 | 2 (1.0%) | 50 (25.0%) |
| Q4 | 37 (18.5%) | 50 (25.0%) |
| Q5 (wealthiest) | 158 (79.0%) | 41 (20.5%) |
| **Covered by health insurance** |  |  |
| Yes | 73 (36.5%) | 200 (100%) |
| **Type of health insurance (if covered), may select more than one if applicable** |  |  |
| National Health Insurance (NHI) | 0 (0%) | 198 (99%) |
| Civil servant | 29 (39.7%) | 0 (0%) |
| Social security | 40 (54.8%) | 0 (0%) |
| Community based health | 0 (0%) | 0 (0%) |
| Health equity fund | 0 (0%) | 0 (0%) |
| Free maternal child health | 1 (1.4%) | 0 (0%) |
| Private health | 5 (6.8%) | 2 (1%) |
| Other | 1 (1.4%) | 0 (0%) |

### Inpatient management and health outcomes

Table 2 summarises the inpatient management and health outcomes of the participants. The length of hospital stay was longer at NCH: median 6 days (IQR 4-7 days) compared to SPH: median 4 days (IQR 3-5 days). The most common primary clinical diagnosis at both hospitals was lower respiratory tract infection or pneumonia (NCH 62.3%, SPH 35.5%). 10.2% of participants from NCH went home before completing treatment, compared to 3.0% from SPH. Across both hospitals, five participants died whilst in hospital, with a further eight participants dying after leaving hospital.

**Table 2.** Clinical diagnoses, inpatient management and health outcomes.

|  | **National Children's**<br>**Hospital N (%)**<br>N=199 | **Salavan Provincial**<br>**Hospital N (%)**<br>N=200 |
| --- | --- | --- |
| **Ward admitted to** |  |  |
| Pediatric General Medicine | 136 (68.4%) | 154 (77.0%) |
| Pediatric Intensive Care Unit (ICU) | 63 (31.7%) | 46 (23.0%) |
| **Length of hospital admission, days** |  |  |
| Median (IQR) | 6 (4-7) | 4 (3-5) |
| **Primary clinical diagnosis** |  |  |
| Lower respiratory tract infection (pneumonia) | 124 (62.3%) | 71 (35.5%) |
| Upper respiratory tract infection | 0 (0.0%) | 43 (21.5%) |
| Acute gastroenteritis and dysentery | 18 (9.0%) | 16 (8.0%) |
| Dengue | 13 (6.5%) | 23 (11.5%) |
| Other infection | 10 (5.1%) | 15 (7.5%) |
| Febrile seizure | 17 (8.5%) | 6 (3.0%) |
| Beriberi | 1 (0.5%) | 7 (3.5%) |
| Other | 16 (8.0%) | 19 (9.5%) |
| **Discharge destination** |  |  |
| Home, treatment completed | 172 (86.4%) | 189 (94.5%) |
| Home, treatment not completed | 20 (10.2%) | 6 (3.0%) |
| Transfer to another health facility | 2 (1.0%) | 4 (2.0%) |
| Deceased | 4 (2.0%) | 1 (0.5%) |
| Other | 1 (0.5%) | 0 (0.0%) |
| **Health outcomes post-discharge\*** |  |  |
| Fully recovered | 146 (76.8%) | 114 (60.3%) |
| Partially recovered, improved since discharge | 33 (17.4%) | 69 (36.5%) |
| Not recovered, same or sicker since discharge | 2 (1.1%) | 2 (1.1%) |
| Deceased (inclusive of deaths in hospital) | 9 (4.7%) | 4 (2.1%) |

\*Complete case analysis: n = 379

#### Out-of-pocket costs

The median household OOP costs from the onset of illness to two months after hospital discharge were summarised by hospital (Table 3). Both mean and median OOP costs were calculated, however due to the skewed distribution of OOP costs and a small number of extreme outliers with very high OOP costs biasing the mean, we used the median OOP costs to compare differences between groups. *A summary of mean household OOP costs and detailed itemised costs are included in S1 and S2 Tables, respectively.* The median total household OOP costs including all direct and indirect costs was over three times higher at NCH: USD290.6 (IQR 206.9 – 422.9) than it was at SPH: USD92.4 (IQR 56.2 – 152.9). Median direct medical OOP costs at NCH was USD128.9 (IQR 70.1 – 218.2) compared with SPH which was USD3.4 (IQR 0.0 – 14.1). There were several outliers, with nine households (six from NCH, three from SPH) incurring more than USD1000 in total OOP costs.

**Table 3.** Out-of-pocket costs (in USD) associated with severe illness, by hospital.

|  | National Children's Hospital<br>N=185* | Salavan Provincial Hospital<br>N=186* |
| --- | --- | --- |
| **Direct medical OOP costs, median (IQR)** | 128.9 (70.1 – 218.2) | 3.4 (0.0 – 14.1) |
| **Direct non-medical OOP costs, median (IQR)** | 65.0 (45.2 – 82.54) | 48.9 (37.9 – 80.3) |
| **Indirect OOP costs, median (IQR)** | 84.8 (45.2 – 132.8) | 25.4 (10.2 – 67.8) |
| **Total OOP (direct medical + direct non-medical costs, median (IQR))** | 195.6 (128.9 – 285.5) | 58.0 (40.1 – 96.1) |
| **Total OOP (direct + indirect costs, median (IQR))** | 290.6 (206.9 – 422.9) | 92.4 (56.2 – 152.9) |

OOP = out-of-pocket; USD = United States Dollar
\*Complete case analysis (ie. Data available for all study visits)

Subgroup analyses (Table 4) revealed that when comparing total OOP costs by wealth subgroups within each hospital, at SPH the wealthiest quintile (Q5) had greater OOP costs than the poorest quintile (Q1): (difference in median OOP costs: USD 69.5, 95% CI 17.3 – 121.7, p=0.009). Other subgroup analyses showed OOP costs were similar by geographical residence, maternal education and ethno-linguistic groups by hospital.

**Table 4.** Comparison of total out-of-pocket costs^^^ (in USD) associated with severe illness, by demographics.

|  | National Children's Hospital (N = 185*) |  |  |  | Salavan Provincial Hospital (N = 186*) |  |  |  |
| --- | --- | --- | --- | --- | --- | --- | --- | --- |
|  | N (%) | OOP costs<sup>^</sup>, median (IQR) | Δ medians (95% C.I.)# | P-value | N (%) | OOP costs<sup>^</sup>, median (IQR) | Δ medians (95% C.I.)# | P-value |
| **Wealth Quintile** |  |  |  |  |  |  |  |  |
| **Q1 (Poorest)** | 0 | N/A | - | - | 17 (9.2%) | 63.9 (45.8, 112.0) | -69.5 (-121.7, -17.3) | 0.009 |
| **Q2** | 0 | N/A | - | - | 34 (18.3%) | 94.4 (48.6, 161.12) | -37.3 (-79.5, 4.9) | 0.082 |
| **Q3** | 1 (0.5%) | 110.2 (110.2, 110.2) | -179.2 (-546.7, 188.3) | 0.337 | 46 (25.3%) | 90.4 (56.8, 152.9) | -40.1 (-79.2, -1.0) | 0.044 |
| **Q4** | 35 (18.9%) | 294.0 (221.9, 466.1) | 4.5 (-64.3, 73.3) | 0.897 | 50 (26.9%) | 83.1 (56.2, 134.3) | -47.5 (-85.9, -9.1) | 0.016 |
| **Q5 (Wealthiest)** | 149 (80.5%) | 289.4 (203.5, 420.9) | Ref | Ref | 39 (21.0%) | 133.4 (72.9, 235.7) | Ref | Ref |
| **Geographical residence** |  |  |  |  |  |  |  |  |
| **Urban** | 165 (89.2%) | 285.5 (198.7, 422.9) | Ref | Ref | 27 (14.5%) | 87.1 (54.27, 216.5) | Ref | Ref |
| **Rural** | 20 (10.8%) | 334.1 (247.1, 401.4) | 48.6 (-42.3, 139.5) | 0.293 | 159 (85.5%) | 93.3 (56.2, 150.4) | 6.6 (-33.9, 46.3) | 0.760 |
| **Maternal education\*** |  |  |  |  |  |  |  |  |
| **None / early childhood** | 7 (3.8%) | 342.9 (212.0, 466.1) | 54.3 (-89.7, 198.2) | 0.458 | 37 (19.9%) | 85.9 (53.7, 129.2) | -7.3 (-47.5, 32.8) | 0.718 |
| **Primary** | 20 (10.8%) | 298.6 (235.3, 379.8) | 14.9 (-73.7, 103.4) | 0.741 | 58 (31.2%) | 102.3 (48.6, 183.7) | 13.8 (-20.9, 48.6) | 0.433 |
| **Secondary or higher** | 154 (83.3%) | 287.0 (203.6, 422.9) | Ref | Ref | 82 (44.1%) | 92.7 (61.9, 188.2) | Ref | Ref |
| **Ethno-linguistic group** |  |  |  |  |  |  |  |  |
| **Lao-Tai** | 162 (87.6%) | 292.6 (208.0, 444.2) | Ref | Ref | 158 (85.0%) | 91.3 (55.4, 148.7) | Ref | Ref |
| **Mon-Khmer** | 5 (2.7%) | 212.0 (147.8, 221.9) | -81.7 (-254.9, 91.5) | 0.353 | 25 (13.4%) | 96.1 (56.8, 183.7) | 4.0 (-36.3, 44.2) | 0.846 |
| **Hmong-Mien** | 16 (8.6%) | 292.3 (187.0, 436.8) | 0.3 (-99.7, 100.3) | 0.996 | 0 | N/A | N/A | N/A |
| **Chinese-Tibetan** | 1 (0.5%) | 248.2 (248.2, 248.2) | -45.5 (-428.2, 337.2) | 0.815 | 0 | N/A | N/A | N/A |
| **Other** | 1 (0.5%) | 313.7 (313.7, 313.7) | 20.1 (-362.6, 402.8) | 0.918 | 3 (1.6%) | 97.8 (65.6, 199.0) | 5.6 (-103.3, 114.6) | 0.919 |

<sup>^</sup>OOP costs = total (direct + indirect) costs associated with severe illness over whole study period
\*Complete case analysis (data available for all study visits). NB for maternal education group – 13 missing values
#Quantile regression analysis to compare differences in median OOP costs across subgroups

#### Catastrophic health expenditures and impoverishment

Table 5 summarises the combined direct and indirect costs calculated as a percentage of household expenditures. At both hospitals, the total OOP costs consumed more than half of the two-monthly household expenditure (NCH: 55.9%, 95% CI 47.3 – 64.4; SPH: 53.3%, 95% CI 37.2 – 69.5). The total OOP costs as a percentage of annual household expenditures was 9.3% (95% CI 7.9 – 10.7) at NCH and 8.9% (95% CI 6.2 – 11.6) at SPH. There were differences between the two hospitals when only direct medical costs were considered, but these differences were less when direct non-medical costs and indirect costs were included in the OOP calculation. Similar results were found when OOP costs were calculated as a percentage of capacity-to-pay.

**Table 5.** Total out-of-pocket costs as a percentage of household expenditure, by hospital.

|  | National Children's Hospital<br>N=183 | Salavan Provincial Hospital<br>N=186 |
| --- | --- | --- |
| **OOP costs as a percentage of two monthly HE – % (95% C.I.)** |  |  |
| **Direct medical costs** | 30.1% (23.8 - 36.4) | 10.6% (0.0 - 22.7) |
| **Direct (medical + non-medical) costs** | 40.7% (33.5 - 47.9) | 38.0% (22.6 - 53.3) |
| **Direct + indirect costs** | 55.9% (47.3 - 64.4) | 53.3% (37.2 - 69.5) |
| **OOP costs as a percentage of two monthly CTP – % (95% C.I.)** |  |  |
| **Direct medical costs** | 35.3% (28.3 - 42.4) | 12.2% (0.0 - 26.4) |
| **Direct (medical + non-medical) costs** | 47.8% (39.8 - 55.9) | 43.2% (25.1 - 61.3) |
| **Direct + indirect costs** | 65.6% (56.0 - 75.2) | 60.4% (41.4 - 79.4) |
| **OOP costs as a percentage of annual HE – % (95% C.I.)** |  |  |
| **Direct medical costs** | 5.0% (4.0 - 6.1) | 1.8% (0.0 - 3.8) |
| **Direct (medical + non-medical) costs** | 6.8% (5.6 - 8.0) | 6.3% (3.8 - 8.9) |
| **Direct + indirect costs** | 9.3% (7.9 - 10.7) | 8.9% (6.2 - 11.6) |
| **OOP costs as a percentage of annual CTP – % (95% C.I.)** |  |  |
| **Direct medical costs** | 5.9% (4.7 - 7.1) | 2.0% (0.0 - 4.4) |
| **Direct (medical + non-medical) costs** | 8.0% (6.6 - 9.3) | 7.2% (4.2 - 10.2) |
| **Direct + indirect costs** | 10.9% (9.3 - 12.5) | 10.1% (6.9 - 13.2) |

OOP = out-of-pocket; HE = household expenditures; CTP = capacity to pay

When comparing CHE rates between the two hospitals, NCH had higher rates of estimated short-term financial hardship (over two months), for all OOP cost categories (Fig 2A and 2B). Estimated longer-term financial impact (over one year) only saw a large difference between the two hospitals when using direct medical costs at a threshold of 10% of annual household expenditures (NCH: 18.5%, 95% CI 13.1 – 23.9; SPH: 8.0%, 95% CI 4.2 – 11.7) with minimal differences at threshold of 40% annual capacity-to-pay (Fig 2C and 2D). *S3 Table provides more detailed results of CHE rates by hospital*.

**Fig 2.**
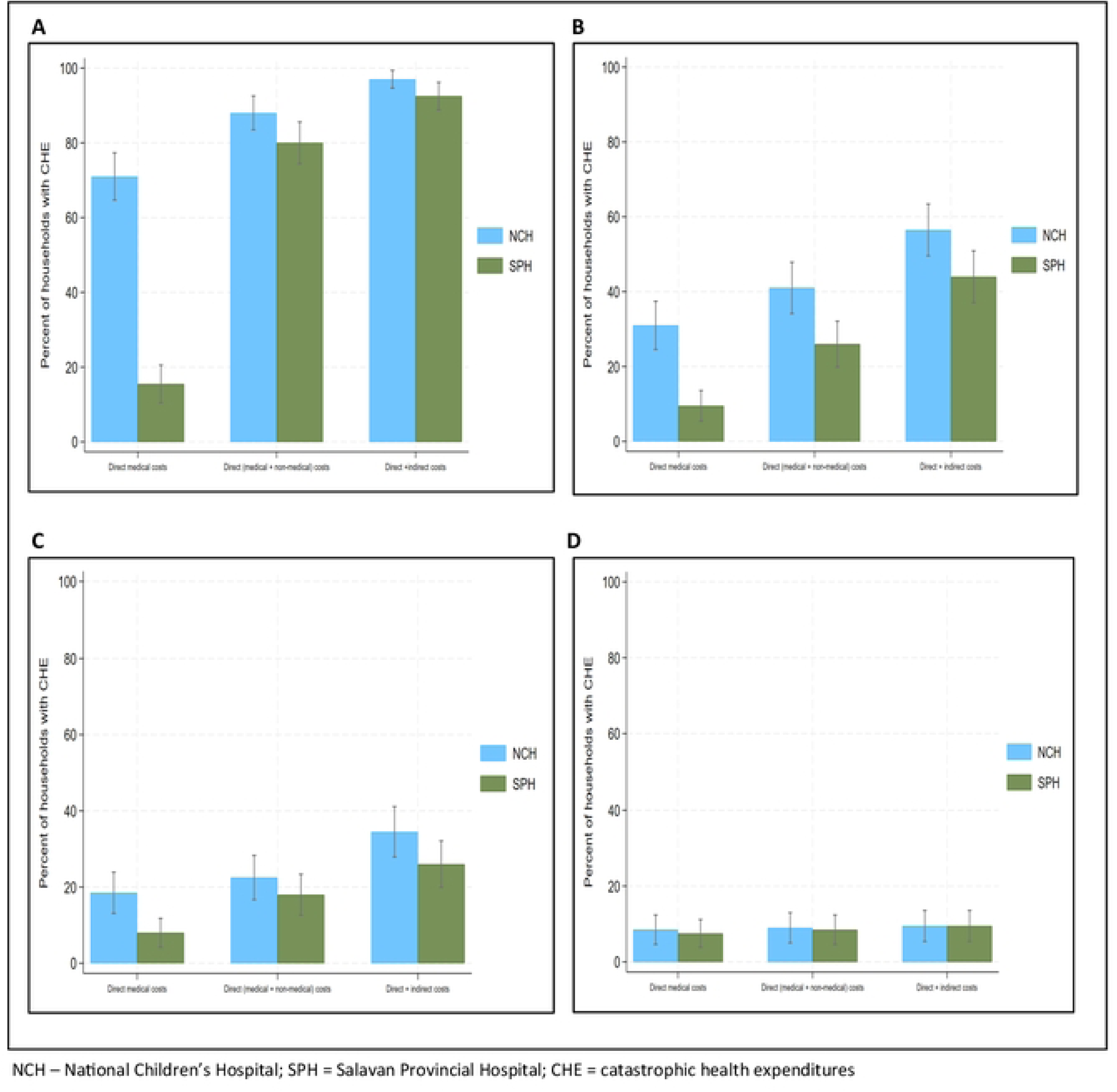
Catastrophic health expenditure rates due to severe illness. A. Threshold: 10% of two-monthly household expenditures B. Threshold: 40% of two-monthly capacity to pay C. Threshold: 10% of annual household expenditures D. Threshold: 40% of annual capacity to pay

Fig 3 shows the poverty headcount associated with the OOP costs by hospital, based on both international and national poverty lines. From enrolment to two months post-discharge, less than five NCH households were impoverished due to OOP costs at both poverty thresholds (Fig 3A and 3C). Based on the international poverty line, 32 (16%) households from SPH were already in poverty at enrolment, with 12 (6%) due to OOP costs before hospital admission. At two months post-discharge, 19 (10.2%) SPH households remained in poverty (Fig 3B). Using the national poverty line, there were higher impoverishment rates from SPH, 69 (34.5%) households at enrolment and 45 (24.1%) at two months post-discharge, with 28 (15%) households impoverished due to OOP costs (Fig 3D). *S4 Table provides more detailed results of impoverishment rates by hospital*.

**Fig 3.**
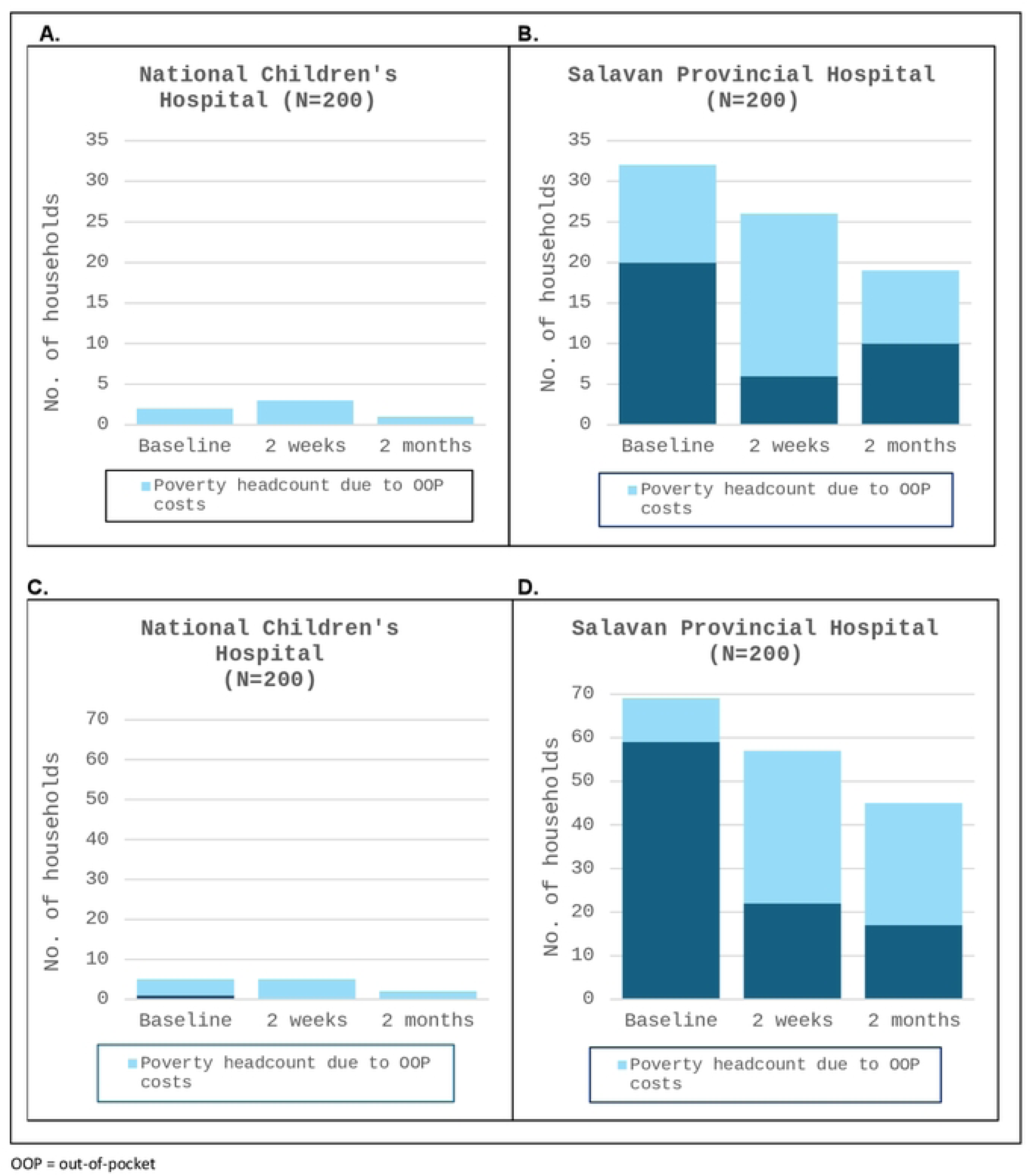
Poverty headcount associated with direct (medical & non-medical) out-of-pocket costs. A. National Children’s Hospital – threshold: International poverty line ($USD 2.15/person/day at 2017 PPP) B. Salavan Provincial Hospital – threshold: International poverty line ($USD 2.15/person/day at 2017 PPP) C. National Children’s Hospital – threshold: Laos national poverty line ($USD 22.00/person/month) D. Salavan Provincial Hospital – threshold: Laos national poverty line ($USD 22.00/person/month)

Tables 6A and 6B summarise the CHE rates by demographic groups at each hospital, with risk ratios calculated across the subgroups. At NCH, higher rates of CHE based on threshold of 10% of annual household expenditures were seen in households from the second poorest quintile (Q2) compared to the wealthiest quintile (Q5) (RR 6.6, 95% CI 4.5 – 9.5), those living in rural versus urban areas (RR 2.4, 95% CI 1.4 – 4.1) and with no formal maternal education compared to secondary education or higher (RR 2.6, 95% CI 1.2 – 5.5). At SPH, higher rates of CHE were similarly seen from the poorest quintile (RR 4.9, 95% CI 1.7 – 13.7), and with no formal maternal education versus secondary education (RR 4.6, 95% CI 1.9 – 11.1), with some evidence of higher CHE rates in rural compared to urban areas (RR 1.9, 95% CI 0.6 – 5.7). Ethnolinguistic group analysis found higher CHE rates amongst the Mon-Khmer group (RR 3.0, 95% CI 1.7 – 5.4) compared to the Lao-Tai group. Refer to tables 6A and 6B for further results of CHE rates across subgroups using other thresholds.

**Table 6A.**
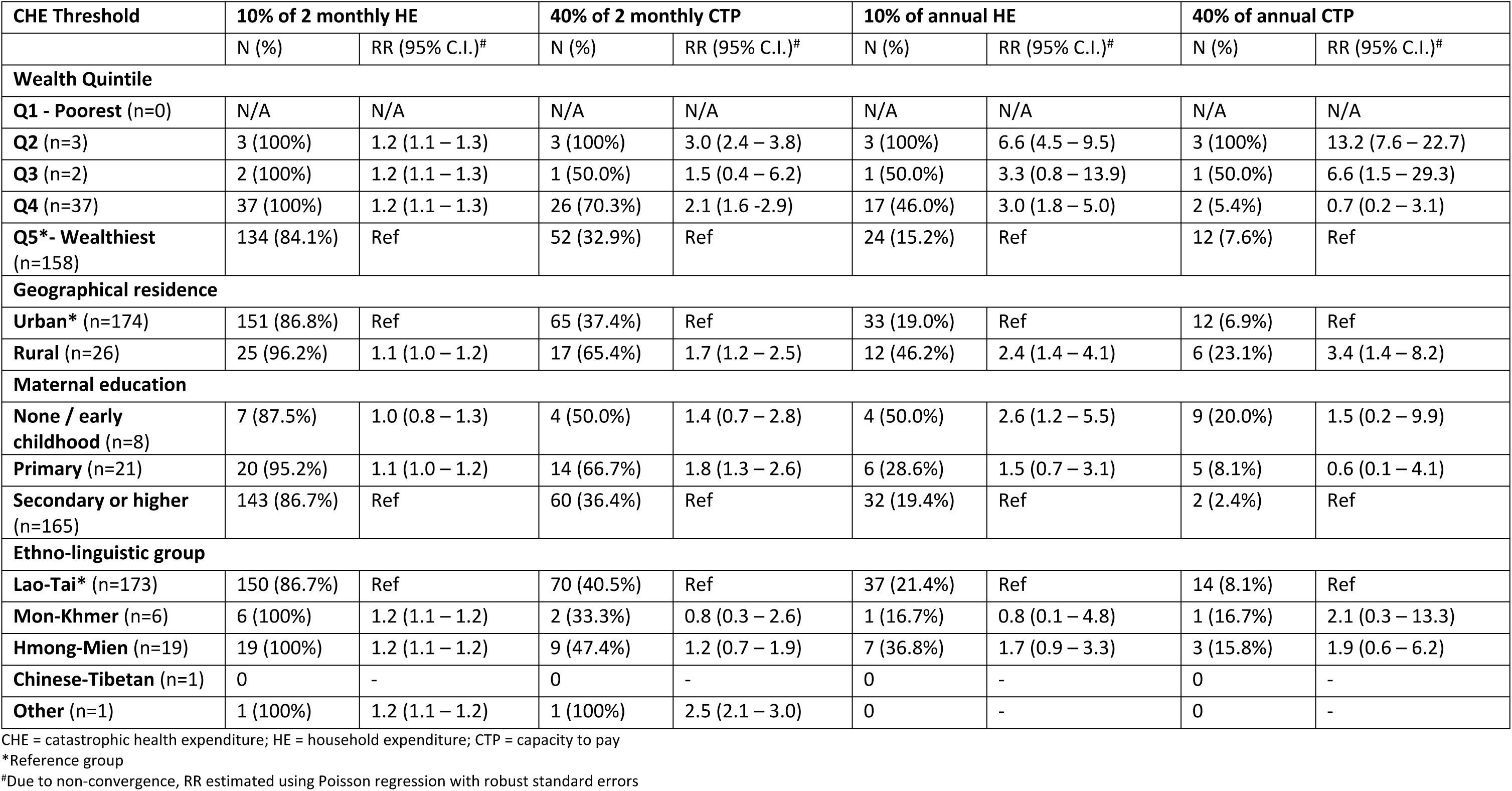
Catastrophic health expenditure rates based on direct (medical + non-medical) costs, by demographics at National Children’s Hospital.

| CHE Threshold | 10% of 2 monthly HE |  | 40% of 2 monthly CTP |  | 10% of annual HE |  | 40% of annual CTP |  |
| --- | --- | --- | --- | --- | --- | --- | --- | --- |
|  | N (%) | RR (95% C.I.)<sup>#</sup> | N (%) | RR (95% C.I.)<sup>#</sup> | N (%) | RR (95% C.I.)<sup>#</sup> | N (%) | RR (95% C.I.)<sup>#</sup> |
| **Wealth Quintile** |  |  |  |  |  |  |  |  |
| **Q1 - Poorest** (n=0) | N/A | N/A | N/A | N/A | N/A | N/A | N/A | N/A |
| **Q2** (n=3) | 3 (100%) | 1.2 (1.1 – 1.3) | 3 (100%) | 3.0 (2.4 – 3.8) | 3 (100%) | 6.6 (4.5 – 9.5) | 3 (100%) | 13.2 (7.6 – 22.7) |
| **Q3** (n=2) | 2 (100%) | 1.2 (1.1 – 1.3) | 1 (50.0%) | 1.5 (0.4 – 6.2) | 1 (50.0%) | 3.3 (0.8 – 13.9) | 1 (50.0%) | 6.6 (1.5 – 29.3) |
| **Q4** (n=37) | 37 (100%) | 1.2 (1.1 – 1.3) | 26 (70.3%) | 2.1 (1.6 -2.9) | 17 (46.0%) | 3.0 (1.8 – 5.0) | 2 (5.4%) | 0.7 (0.2 – 3.1) |
| **Q5\*- Wealthiest** (n=158) | 134 (84.1%) | Ref | 52 (32.9%) | Ref | 24 (15.2%) | Ref | 12 (7.6%) | Ref |
| **Geographical residence** |  |  |  |  |  |  |  |  |
| **Urban\*** (n=174) | 151 (86.8%) | Ref | 65 (37.4%) | Ref | 33 (19.0%) | Ref | 12 (6.9%) | Ref |
| **Rural** (n=26) | 25 (96.2%) | 1.1 (1.0 – 1.2) | 17 (65.4%) | 1.7 (1.2 – 2.5) | 12 (46.2%) | 2.4 (1.4 – 4.1) | 6 (23.1%) | 3.4 (1.4 – 8.2) |
| **Maternal education** |  |  |  |  |  |  |  |  |
| **None / early childhood** (n=8) | 7 (87.5%) | 1.0 (0.8 – 1.3) | 4 (50.0%) | 1.4 (0.7 – 2.8) | 4 (50.0%) | 2.6 (1.2 – 5.5) | 9 (20.0%) | 1.5 (0.2 – 9.9) |
| **Primary** (n=21) | 20 (95.2%) | 1.1 (1.0 – 1.2) | 14 (66.7%) | 1.8 (1.3 – 2.6) | 6 (28.6%) | 1.5 (0.7 – 3.1) | 5 (8.1%) | 0.6 (0.1 – 4.1) |
| **Secondary or higher** (n=165) | 143 (86.7%) | Ref | 60 (36.4%) | Ref | 32 (19.4%) | Ref | 2 (2.4%) | Ref |
| **Ethno-linguistic group** |  |  |  |  |  |  |  |  |
| **Lao-Tai\*** (n=173) | 150 (86.7%) | Ref | 70 (40.5%) | Ref | 37 (21.4%) | Ref | 14 (8.1%) | Ref |
| **Mon-Khmer** (n=6) | 6 (100%) | 1.2 (1.1 – 1.2) | 2 (33.3%) | 0.8 (0.3 – 2.6) | 1 (16.7%) | 0.8 (0.1 – 4.8) | 1 (16.7%) | 2.1 (0.3 – 13.3) |
| **Hmong-Mien** (n=19) | 19 (100%) | 1.2 (1.1 – 1.2) | 9 (47.4%) | 1.2 (0.7 – 1.9) | 7 (36.8%) | 1.7 (0.9 – 3.3) | 3 (15.8%) | 1.9 (0.6 – 6.2) |
| **Chinese-Tibetan** (n=1) | 0 | - | 0 | - | 0 | - | 0 | - |
| **Other** (n=1) | 1 (100%) | 1.2 (1.1 – 1.2) | 1 (100%) | 2.5 (2.1 – 3.0) | 0 | - | 0 | - |

CHE = catastrophic health expenditure; HE = household expenditure; CTP = capacity to pay
\*Reference group
<sup>#</sup>Due to non-convergence, RR estimated using Poisson regression with robust standard errors

**Table 6B.** Catastrophic health expenditure rates based on direct (medical + non-medical) costs, by demographics at Salavan Provincial Hospital.

| CHE Threshold | 10% of 2 monthly HE |  | 40% of 2 monthly CTP |  | 10% of annual HE |  | 40% of annual CTP |  |
| --- | --- | --- | --- | --- | --- | --- | --- | --- |
|  | N (%) | RR (95% C.I.)<sup>#</sup> | N (%) | RR (95% C.I.)<sup>#</sup> | N (%) | RR (95% C.I.)<sup>#</sup> | N (%) | RR (95% C.I.)<sup>#</sup> |
| **Wealth Quintile** |  |  |  |  |  |  |  |  |
| **Q1 - Poorest** (n=21) | 20 (95.2%) | 1.6 (1.2 – 2.0) | 12 (57.1%) | 3.9 (1.7 – 8.9) | 10 (47.6%) | 4.9 (1.7 – 13.7) | 5 (23.8%) | 4.9 (1.0 – 23.2) |
| **Q2** (n=38) | 35 (92.1%) | 1.5 (1.2 – 2.0) | 14 (36.8%) | 2.5 (1.1 – 5.9) | 12 (31.6%) | 3.2 (1.1 – 9.2) | 4 (10.5%) | 2.2 (0.4 – 11.2) |
| **Q3** (n=50) | 42 (84.0%) | 1.4 (1.1 – 1.8) | 16 (32.0%) | 2.2 (0.9 – 5.1) | 7 (14.0%) | 1.4 (0.1 – 2.6) | 5 (10.0%) | 2.1 (0.4 – 10.1) |
| **Q4** (n=50) | 38 (76.0%) | 1.2 (0.9 – 1.7) | 4 (8.0%) | 0.5 (0.2 – 1.8) | 3 (6.0%) | 0.6 (0.1 – 2.6) | 1 (2.0%) | 0.4 (0.0 – 4.4) |
| **Q5\*- Wealthiest** (n=41) | 25 (61.0%) | Ref | 6 (14.6%) | Ref | 4 (9.8%) | Ref | 2 (4.9%) | Ref |
| **Geographical residence** |  |  |  |  |  |  |  |  |
| **Urban\*** (n=29) | 17 (58.6%) | Ref | 4 (13.8%) | Ref | 3 (10.3%) | Ref | 2 (6.9%) | Ref |
| **Rural** (n=171) | 143 (83.6%) | 1.4 (1.0 – 1.9) | 48 (28.1%) | 2.0 (0.8 – 5.2) | 33 (19.3%) | 1.9 (0.6 – 5.7) | 15 (8.8%) | 1.3 (0.3 – 5.3) |
| **Maternal education** |  |  |  |  |  |  |  |  |
| **None / early childhood** (n=53) | 44 (97.8%) | 1.6 (1.3 – 1.8) | 20 (44.4%) | 2.8 (1.6 – 5.2) | 15 (33.3%) | 4.6 (1.9 – 11.1) | 9 (20.0%) | 8.3 (1.9 – 36.9) |
| **Primary** (n=83) | 55 (88.7%) | 1.4 (1.2 – 1.7) | 17 (27.4%) | 1.7 (0.9 – 3.3) | 14 (22.6%) | 3.1 (1.3 – 7.7) | 5 (8.1%) | 3.4 (0.7 – 16.8) |
| **Secondary or higher** (n=248) | 52 (62.7%) | Ref | 13 (15.7%) | Ref | 6 (7.2%) | Ref | 2 (2.4%) | Ref |
| **Ethno-linguistic group** |  |  |  |  |  |  |  |  |
| **Lao-Tai\*** (n=168) | 129 (76.8%) | Ref | 37 (22.0%) | Ref | 23 (13.7%) | Ref | 11 (6.6%) | Ref |
| **Mon-Khmer** (n=29) | 28 (96.6%) | 1.3 (1.1 – 1.4) | 14 (48.3%) | 2.2 (1.4 – 3.5) | 12 (41.4%) | 3.0 (1.7 – 5.4) | 6 (20.7%) | 3.2 (1.3 – 7.9) |
| **Hmong-Mien** (n=0) | N/A | N/A | N/A | N/A | N/A | N/A | N/A | N/A |
| **Chinese-Tibetan** (n=-0) | N/A | N/A | N/A | N/A | N/A | N/A | N/A | N/A |
| **Other** (n=3) | 3 (100%) | 1.3 (1.2 – 1.4) | 1 (33.3%) | 1.5 (0.3 – 7.7) | 1 (33.3%) | 2.4 (0.5 – 12.7) | 0 | - |

CHE = catastrophic health expenditure; HE = household expenditure; CTP = capacity to pay
\*Reference group
<sup>#</sup>Due to non-convergence, RR estimated using Poisson regression with robust standard errors

Risk ratios were also calculated across demographic groups for impoverishment rates. At SPH, higher impoverishment rates based on the national poverty line, were seen in the poorest (Q1) versus wealthiest quintile (Q5) (RR 2.6, 95% CI 1.3 – 5.4), households with no formal maternal education compared to secondary education or higher (RR 7.4, 95% CI 1.1 – 51.9) and the Mon-Khmer versus Lao-Tai ethnic groups (RR 2.4, 95% C.I. 1.4 – 4.1). At NCH, findings were limited due to overall low numbers of impoverishment. *Details of these results are included in S5 and S6 Tables*.

### Financial coping strategies

The OOP costs were calculated after health insurance rebates (where applicable) were accounted for. Further financial coping strategies that households reported to help cover OOP costs related to the severe illness at discharge from hospital included: use of savings (overall 394/400 = 98.5%), borrowing money from relatives or friends (NCH 17.9%, SPH 14.5%), reducing household expenses (NCH 3.1%, SPH 0%), selling assets (two households), borrowing money from the bank (two households) and delaying plans (one household). *Refer to S7 Table for a summary of coping strategies reported at each study visit*.

## Discussion

Our study found that in a secondary level, provincial hospital in Lao PDR where the NHI is available, direct medical costs for inpatient treatment were lower, with households paying a median of USD3.4 compared to households using a tertiary level, central hospital without NHI, who paid a median of USD128.9 in direct medical costs. However, total household OOP costs (direct and indirect) related to severe childhood illnesses were high overall and resulted in substantial financial distress in households from both SPH and NCH. CHE (direct OOP costs >10% of annual household expenditures) rates in our study (NCH 22.5%, SPH 18.0%) were higher than globally reported CHE rates of 13.5%.[11] Based on the international poverty line, impoverishment rates in our study as a result of direct OOP costs were higher at SPH (10.2% - 16.0%) than NCH (0.5% - 1.5%). This is in comparison to global impoverishment rates from direct OOP costs of 4.4%.[11]

Total household OOP costs at NCH were three times higher than at SPH. By differentiating the differences in the types of OOP costs, higher OOP costs at NCH households were likely due to patients at NCH on average having a higher ICU admission rate with longer hospital stays than SPH patients, resulting in higher direct medical costs, as well as the NHI covering almost all the direct medical costs at SPH. Tertiary hospitals compared to secondary level hospitals are also generally more expensive. They have higher bed costs, treat more complex cases, with access to more advanced equipment and diagnostics with higher associated costs.[32, 33] Furthermore, at NCH where the primary income earners were mainly manual labourers or service providers, households also had higher estimated incomes than at SPH, where the household heads were predominantly farmers. This resulted in higher indirect costs at NCH for estimated loss of productivity.

Despite NCH having higher OOP costs than SPH, the CHE rates were similar between the two hospitals and impoverishment rates were conversely higher at SPH. The denominators in CHE and impoverishment calculations were dependent on estimated household consumption. The expenses collected including food, housing, utility bills, transportation, education, shopping and recreation are a reflection of local living costs and spending patterns, with urban areas, particularly major cities, generally having higher living costs than rural areas. In Lao PDR, where a large proportion of the wealthiest population live in Vientiane Capital, recent national data showed that the wealthiest quintile consume seven times more on average per capita than the poorest quintile, and rural areas spend more of their household budget on food compared to urban areas (68% versus 53%).[14] These contrasts in demographics and spending patterns can begin to explain the factors influencing our study differences in household expenditure, with NCH having over double the household consumption than SPH, inadvertently resulting in similar CHE rates. Our study however, is not representative of all the demographics in Lao PDR and more studies are required in other settings to further evaluate the complex nature of health related financial burdens on households as well as the impact of the NHI.

CHE and impoverishment rates found in our study were high compared with other South-East Asian countries. A study by Wang et al, looking at rates of financial protection in eight countries in South- East Asia (Bangladesh, Bhutan, India, Maldives, Nepal, Sri Lanka, Thailand and Timor-Leste), found CHE rates (direct medical OOP costs >10% of annual household expenditures) ranging from 1.9% (Thailand) to 19.9% (Maldives). Impoverishment rates using the international poverty line ranged from 0% (Thailand) to 4.2% (India).[6] Based on the same definitions and thresholds, our study had a CHE rate of 13.3% and impoverishment rates of 2.7% - 5.9%. Thailand, which neighbours Lao PDR, is seen as an example where government investment in health and a well implemented NHI scheme has translated to lower financial hardship rates on par with high income countries.[34] Lao PDR is yet to emulate similar results from their NHI scheme, with challenges of insufficient budget and low revenues from copayments, operational issues and continuing government health expenditures negatively impacted by COVID-19.[22]

Large equity disparities between demographic subgroups were found in our study with higher financial hardship rates occurring in the poorest households, those living in rural areas, households with low levels of maternal education and ethnic minority groups. Similar trends of inequity have been shown in other LMIC studies assessing the financial burden for sick children requiring health care. A Kenyan study found that CHE rates (direct medical costs >40% of capacity-to-pay) occurred almost four times as often in lower socio-economic households than higher ones.[35] A study of sick children in Bangladesh showed that households with mothers who had no formal education needed external financial assistance to fund OOP costs three times more often than households with maternal secondary education levels or higher.[36] Inequitable financial hardship however, are not limited to LMICs. A systematic review on the determinants of CHE found similar socio-economic disparities in CHE rates to our study, across low, middle and high income countries, with health insurance associated with lower CHE rates in only 9/17 studies.[37] Health equity is a key feature of UHC but remains a global challenge. Health insurance plays a role towards improving health equity, but further interventions are required to address all the factors that impact socio-economic disparities in CHE and impoverishment.

The NHI scheme aims to improve equitable access to health care and reduce unmet medical needs. Studies in Lao PDR prior to NHI introduction, found conversely lower CHE rates in poorer households compared to wealthier ones. These studies suggested that this was a reflection of the poor being less likely to seek medical care rather than being financially protected. Difficult access and high costs of health treatment were cited as reasons for not seeking health care.[28, 38] Since the introduction of the NHI in Lao PDR, there is some evidence that the NHI has increased health service utilisation for the poorest households.[15, 39] There was also indication of this in our study where participants from SPH where the NHI is available, were relatively equally distributed across wealth quintiles. In NCH where NHI is not available, participants were almost exclusively from the two wealthiest quintiles. This suggests poorer households in the Capital City with severely sick children either sought care from other health centres or did not receive health care at all. Our study also had a small proportion of participants from ethnic minority groups at both hospitals with most (85.3%) from the Lao-Tai ethno-linguistic group, compared to a national average of 62.4%.[21] This may be due to our limited sample not being representative of the country and inclusion criteria of parents of participants required to speak the Lao language, but it may also reflect cultural barriers to accessing health care. Studies from neighbouring Asian countries have shown reasons for low uptake of healthcare services by ethnic minority groups included being from poorer households, living in remote areas with limited transportation, having lower health literacy levels, and culturally inappropriate practices from medical personnel.[40, 41] The NHI aims to mitigate some of these barriers by providing financial incentives in addition to free health care, with travel and food vouchers for the poor and those living in remote areas. Despite this, participants from minority groups in our study were low. More research extending to other provinces where the NHI is available, is required to further assess the impact of NHI on equitable access to care. Further evaluation is also needed into the reasons for lower participant numbers from ethnic minority groups and poorer households in NCH to ensure these vulnerable populations in the Capital City are not overlooked in any health programs and policy reforms.

Definitions for financial protection in the literature are inconsistent, requiring careful interpretation and comparison of results. In our study we calculated and described OOP costs at two hospitals using both direct and indirect expenses. This was to identify the differences in types of OOP costs between the two hospitals, but also to recognise the contribution of indirect costs towards financial hardship on households. The SDGs use direct (medical and non-medical) OOP expenses, to calculate CHE and impoverishment rates which ignores the impact of indirect costs on households. In the case of children, the need for an accompanying caregiver may have a significant financial burden if the caregiver is unable to make any income for the duration they are caring for their sick child.[2] Our study found that adding in indirect costs increased the total OOP expenses by 50% – 60%. Similar findings were seen in a study in Malaysia evaluating the financial impact of childhood gastroenteritis requiring hospitalisation which showed indirect costs contributed 44% – 74% to total OOP costs and increased CHE rates at one of the hospitals from one third to 88% of households, compared with direct OOP costs alone.[42] A further hidden impact of indirect costs are the missed healthcare opportunities where families decide to not seek care or leave hospital early due to inaffordability with likely negative health impacts. In our study, 26/400 (6.6%) patients were discharged early without completing medical treatment, four of whom died during the follow-up period.

There are also variable thresholds to define CHE and impoverishment. The SDGs use total household income or expenditures as the denominator to calculate CHE. Previous studies however, have shown that using the household “capacity-to-pay” as the denominator yields a more accurate picture of CHE for poorer households, as they spend a larger proportion of their income on subsistence items, with little left over for discretionary expenses.[2, 4, 5, 25–27] In Lao PDR where much of the country relies on informal economic activity and subsistence farming, using capacity-to-pay may better reflect the financial burden of OOP costs in vulnerable populations.[15, 43] This was demonstrated in our study with larger equity gaps in CHE rates when using capacity-to-pay compared with using household expenditure as the denominator, between the poorest and wealthiest quintiles, between rural and urban areas and between those with no education and higher education levels. Our study included both international and national poverty lines to calculate health-related impoverishment rates. The international poverty line can be used to compare extreme poverty levels across countries. However, the Lao PDR national poverty line and the items used to calculate it, better reflects the living standards and spending patterns within the country, allowing more accurate comparisons across subpopulations.[14] In our study, poverty headcounts using the national poverty line showed larger inequity gaps negatively affecting, the poor, those living those living outside the Capital City and in rural areas, with lower maternal education levels and ethnic minority groups, than when using the international poverty line.

A limitation of the current metrics used to calculate and identify rates of financial protection is that they do not consider how households pay for OOP health expenses, known as “financial coping mechanisms”. In LMICs, including Lao PDR, it is common for families to borrow money, use savings or sell assets and land to pay for OOP health expenditures, with previous studies showing rural households and those of lower income more likely to use multiple coping strategies and/or borrow larger amounts of money.[5, 44–46] The impact of financial coping mechanisms can continue well pass the physical health outcomes. Households may struggle to repay loans and resort to cutting- back on subsistence spending, exposing them to further stressors.[46] A Bangladeshi study on children admitted into hospital for pneumonia, found that of the families who borrowed money to pay for OOP expenses, almost half anticipated needing more than one year to repay the loan.[45] Our study showed there was lingering financial burden two months after being discharged from hospital, with almost half of the households reporting still using savings to pay for OOP expenses and around 3% of households yet to pay back money borrowed from relatives or friends. Longer follow-up periods are required to determine how long these financial burdens persist.

Our study had several limitations. Data was collected primarily from self-report, potentially leading to recall bias. We tried to reduce this by limiting the recall period to two weeks, in line with the LECS6 methodology and providing diaries for households to document expenses inbetween study visits. Our study only included patients from two hospitals, and are not representative of the entire population. Non Lao speaking parents were excluded from the study which may have omitted more disadvantaged ethnic minority groups and potentially biased the cohort. We did not have an equal distribution across wealth quintiles from NCH and therefore could not assess the financial impact of health care costs on poorer groups in the Capital City, likely underestimating CHE and impoverishment rates. Further study is required to understand barriers to accessing care and how the NHI scheme has impacted this, with focus on highly vulnerable groups not eligible for NHI being considered in any restructuring of the NHI scheme.

## Conclusion

Severe childhood illnesses can result in high OOP costs leading to financial hardship for households. This is the first study since the introduction of the NHI scheme, to assess and describe the financial burden on households in Lao PDR when sick children are admitted into hospital. Our study shows that while direct medical OOP expenses for hospitalised children are relatively low where NHI is available, CHE and impoverishment rates remain high with inequitable distribution towards the poor, those living in rural areas, with lower maternal education levels and ethnic minority groups. Further investment in interventions beyond the NHI is needed, including evaluation of their impact, to prevent severe childhood illnesses and reduce non-medical related costs of illnesses that contribute to financial hardship. Additional study is required to explore underlying causes of disparities among disadvantaged groups including those in the Capital City not eligible for NHI. These populations need further financial protection and support accessing health services to reduce health inequities and overall financial burden on households and in Lao PDR.

## Data Availability

De-identified, individual participant data that underlie the results reported in this article can be made available upon request and approval through The Royal Children's Hospital, Human Research Ethics Committee, who will review the proposed use of data. The individual participant data have not been made publicly available due to the sensitive nature of the data (including but not limited to participants being minors, medical treatment, household income). Participant consent obtained at the time of the study also did not include public sharing of data.

## Acknowledgments

We would like to thank the families for participating in the study, the staff at NCH and SPH for their support with the study and for the infrastructure assistance of LOMWRU. We would like to acknowledge the following members for their contribution to this project: Dr Koukeo Phommasone for support with site visits to NCH and SPH, including communication and translation with hospital staff; Dr Keooudomphone Vilivong and Dr Cindy Chu for their clinical support and management of the research team; and Athirat Black, Darren Ong and Lakshi Starks for their administrative support. And thank you to A/Prof Claire von Mollendorf and Prof Steve Graham for their contribution towards the final manuscript.

## Supporting information

**S1 Table: Mean out-of-pocket costs (in USD) associated with severe illness, by hospital**

USD = United States Dollar, OOP = out-of-pocket

*Complete case analysis (ie. Data available for all study visits)

**S2 Table: Itemised costs contributing to total out-of-pocket costs (in USD) associated with severe illness by hospital**

USD = United States Dollar

*Visit 1 = at enrolment during hospital admission; Visit 2 = at hospital discharge; Visit 3 = 2 weeks post hospital discharge; Visit 4 = 2 months post hospital discharge

**S3 Table: Catastrophic health expenditure rates associated with severe illness, by hospital**

HE = household expenditures; CTP = capacity to pay

**S4 Table: Impoverishment rates based on direct (medical & non-medical) OOP costs associated with severe illness, by hospital**

USD = United States Dollar; LAK = Lao Kip

**S5 Table. Impoverishment rates based on direct (medical + non-medical) costs, by demographics at National Children’s Hospital**

USD = United States Dollar; LAK = Lao Kip

*Reference group

^#^Due to non-convergence, RR estimated using Poisson regression with robust standard errors

**S6 Table: Impoverishment rates based on direct (medical + non-medical) costs, by demographics at Salavan Provincial Hospital**

USD = United States Dollar; LAK = Lao Kip

*Reference group

^#^Due to non-convergence, RR estimated using Poisson regression with robust standard errors

**S7 Table: Reported financial coping strategies to pay for out-of-pocket costs associated with severe illness by hospital**

*Visit 1 = at enrolment during hospital admission; Visit 2 = at hospital discharge; Visit 3 = 2 weeks post hospital discharge; Visit 4 = 2 months post hospital discharge

